# Health care seeking behaviors in Puerperal sepsis in rural Sindh, Pakistan: a qualitative study

**DOI:** 10.1101/2020.08.13.20174151

**Authors:** Shabina Ariff, Ubaidullah Khan, Ali Turab, Sumra Kureishy, Farrukh Raza, Farhana Tabassum, Fatima Mir, Amnesty LeFevre, Linda A. Bartlett, Atif Habib, Sajid Bashir Soofi, Zulfiqar A Bhutta

## Abstract

**Background:** Puerperal sepsis (PS) is one of the major causes of maternal death, contributing to 26 000 deaths per year in developing countries. Early recognition and treatment are essential to managing PS, but numerous social, cultural and technical barriers prevent or delay access to care and necessary medical attention. Through this qualitative study, we identified barriers to care seeking for puerperal sepsis among recently delivered women in Matiari, Pakistan.

**Methods:** We conducted 20 in-depth interviews among recently delivered women with and without sepsis and their family members. Key informant interviews were conducted with 14 healthcare providers and traditional birth attendants. The themes used for content analysis were knowledge of danger signs, factors affecting care seeking and local treatment practices for postpartum sepsis.

**Results:** Recently delivered women, their family members and traditional birth attendants were unaware of the word PS or the local translated term for PS. However, they were familiar with most of the individual symptoms associated with PS. Healthcare providers were aware of the condition and the associated symptoms. The healthcare providers understanding of the seriousness of PS was directly proportional their age and clinical experience. The most common barriers to care seeking was the division of labor within the household, obtaining permission from the primary decision maker, access to transportation, lack of financial resources and support from family members.

**Discussion:** In rural Pakistan, limited knowledge of PS, division of labor within the household, obtaining permission from primary decision maker and access to transportation and financial resources were identified as barriers to seeking care among recently delivered women.

**Conclusion:** To improve maternal care seeking behaviors for PS, interventions focusing on increasing knowledge of PS, addressing gender inequality, implementing an affordable community transport service and enhancing TBA’s knowledge and skills to manage PS need to be implemented.

## Background

Puerperal sepsis (PS) is a serious infection of the genital tract contracted by women during or after childbirth or miscarriage [1]. It is a common cause of maternal morbidity and mortality worldwide [2]. According to WHO, in developing countries, PS is the third leading cause of maternal mortality, contributing to 26 000 deaths per year [1]. In 2014, global estimates suggested over a period of ten years 260 000 maternal deaths were attributed to PS, with approximately 90% of these deaths occurring in Sub-Saharan Africa and Southern Asia [1]. In Pakistan, PS is the third leading cause of maternal mortality after hemorrhage and eclampsia [4]. Among Pakistani women, it is estimated to have contributed to 16.3% of maternal deaths in 2002 [5].

The 2013 Pakistan Demographic and Health Survey (PDHS) provided limited data on maternal care seeking behaviors during the postpartum period [6]. Nonetheless, the survey identified some problems in accessing healthcare for women, such as asking permission from family, arranging money, the distance to the facility, access to transportation and the presence of a companion. A study conducted in urban Pakistan further identified household socioeconomic status and perception of symptom severity as barriers to care seeking [5,7].

However, there is limited literature on the barriers to care seeking among women with PS. The research available provides insight into overall postpartum morbidity and associated barriers to care seeking [12, 13]. Thaddeus and his colleagues developed a Three Delays Model to provide an explanation of the factors that prevent mothers from seeking care at health facilities in developing countries [8–10]. These factors were categorized into three groups; socioeconomic and cultural characteristics, accessibility of health facilities and quality of care. Researchers have validated this model across many countries and regions [11]. We anticipated that factors of the Three Delays Model would also be present in Pakistan. To overcome the lack of data on care seeking for PS, we conducted a study to examine the barriers and patterns of care seeking behaviors among recently delivered women (RDW) with PS in rural Pakistan, utilizing the Three Delays Model for analyses.

## Methods

### Study Design

We conducted a qualitative study focused on identifying the barriers to care seeking for PS among RDW in Matiari, Pakistan.

### Study Setting

This study was conducted in Matiari, a rural district situated in the southern Sindh province of Pakistan (figure 1). The population of Matiari is approximately 0.6 million. To recruit eligible participants, other projects participant datasets were used to identify RDW with or without puerperal sepsis in the community. The traditional birth attendants (TBA) and healthcare providers were randomly selected to participate in the study.

### Study Participants

In September 2012, 20 in-depth interviews (IDI) were conducted with the RDW with or without PS and their family members. Key informant interviews (KII) were conducted with 14 healthcare providers (Table 1). The study participants were selected from health facilities and within the community. From the community setting, 7 RDW without PS, 6 family members of RDW and 8 healthcare providers were interviewed at their households. From the health facilities, 4 RDW with PS seeking care, 3 family members of RDW and 6 healthcare providers with experience in managing and treating PS were interviewed at the Hyderabad Civil Hospital. Prior to conducting the interviews, informed consent was obtained from all participants. Participants were selected irrespective of age, socioeconomic status, and occupational, educational and religious backgrounds.

**Table 1:** Participants’ Classification, Interview Type & Frequency.

| Participants' Classification | Interview Type | Frequency |
| --- | --- | --- |
| Healthcare Providers in Health Facilities | Key Informant Interviews | 6 |
| Hospitalized RDW with PS | In-depth Interviews | 4 |
| Family member of Hospitalized RDW with PS | In-depth Interviews | 3 |
| RDW with or without PS | In-depth Interviews | 7 |
| Senior Female of the Household | In-depth Interviews | 6 |
| Healthcare Providers in Community (TBA) | Key Informant Interviews | 8 |

### Data Collection and Analysis

Trained research moderators with experience in qualitative data collection conducted the Interviews in Sindhi (the local language spoken in Matiari). The interviews lasted 35-60 minutes and explored participants’ perceptions of the causes, symptoms, symptoms recognition, sources of care, treatment modalities, and healthcare seeking behaviours towards PS. A moderator note taker and observer were present during all Interviews. All interviews were transcribed, translated into English and entered into a computer by the note taker. Transcript reviews were conducted by research officers to ensure quality control.

Content analysis was conducted manually by an independent data analyst. The data analyst conducted transcript reviews and response categorization to identify the themes present within the data. The themes emerged during data review and were not predetermined. The themes identified were knowledge of danger signs, factors affecting care seeking and local treatment practices for postpartum sepsis.

### Ethical Considerations

The Ethics Review Committee (ERC) of Aga Khan University granted approval for the study. Informed consent was obtained from all participants prior to the interviews. The consent form provided Information on study objectives, procedures, and process of confidentiality.

## Results

### Participant Characteristics

In September 2012, a total of 34 interviews were conducted with RDW, family members of RDW and healthcare providers. Of the 34 female participants, 44% were 18-35 years old, 47% were housewives and 70% were Illiterate. There were 14 healthcare providers: 8 TBA and 6 physicians. Table 2 summarizes the demographic characteristics of participants.

**Table 2:** Demographic Characteristics of Participants.

| Characteristics | Study Settings |  | Total (n=34) |
| --- | --- | --- | --- |
|  | Health Facility (n=13) | Community (n= 21) |  |
| Age (Years) |  |  |  |
| 18-35 | 8 | 7 | 15 |
| 36-50 | 3 | 6 | 9 |
| >50 | 2 | 8 | 10 |
| Education |  |  |  |
| Illiterate | 18 | 6 | 24 |
| Class 1-5 | 4 | 0 | 4 |
| Class 6-10 | 0 | 0 | 0 |
| Class 11-15 | 0 | 0 | 0 |
| Class > 16 | 0 | 6 | 6 |
| Occupation |  |  |  |
| Doctor | 0 | 6 | 6 |
| TBA | 8 | 0 | 8 |
| Housewife | 10 | 6 | 16 |
| Others | 0 | 0 | 0 |
| Religion |  |  |  |
| Islam | 18 | 12 | 32 |

### Knowledge of PS

Most participants (RDW, family members of RDW and TBA) were unaware of the word PS or the local translated term for PS. After providing an explanation of PS and related symptoms, participants were still unable to establish or associate any local term with the condition. However, they were able to elaborate on the individual signs and symptoms of PS without associating them directly to the condition. Participants were unaware of PS being a public health concern and contributing to maternal morbidity and mortality. Generally, RDW associated the signs and symptoms of PS with general weakness. Alternatively, TBA had knowledge of the signs and symptoms related to postpartum hemorrhage and retained placenta. They were also very comfortable discussing the obstetric and gynecological conditions.

The physicians interviewed had good knowledge of PS, and the seriousness and impact of the condition on maternal morbidity and mortality. A physician’s understanding of the seriousness of PS was directly proportional the physician’s age and clinical experience. For instance, the younger physicians Interviewed seemed less aware of the impact of PS on maternal mortality. Additionally, there was no consensus on symptoms for diagnosing PS among the physicians. They agreed that maternal antenatal and natal history along with the presence of fever should be considered when diagnosing PS. However, their opinions varied concerning the use of other postpartum complaints (vaginal discharge, bleeding and pelvic tenderness) for diagnosing PS. The physicians also understood that fever may be related to other conditions such as malaria, typhoid and urinary tract infections.

### Patterns of Care Seeking Behaviors for PS

As previously mentioned, RDW were able to relate to the individual symptoms of PS. Fever was the most frequently mentioned symptom of PS among RDW, family members of RDW and TBA. However, the presence of fever alone was not considered crucial by RDW and family members to care seeking. On the other hand, healthcare providers (physicians and TBA) considered fever alone as an important symptom to encourage care seeking. Pain in pelvic region and vaginal discharge were also Identified as important symptoms to prompt care seeking among RDW and family members of RDW. One RDW recounted her experience with care seeking for PS.

> *My baby was born through the normal way by the grace of God. First*, *I was fine but I started having fever soon after delivery. The fever increased everyday but it was only fever so I didn’t go to seek health care; if I start going to doctor for small complaints then who will tend to my children and do household chores. Elder women of my family also assured me that its normal to have fever after delivery. After some days I also started having pain in pelvis. When I noticed there was also foul-smelling water coming from vagina. My mother in law advised me to visit the ‘Shamshad’ the TBA who also delivered my baby. The TBA said fever shouldn’t be a problem after delivery and gave me some medicines. My symptoms worsened the same night and family was thinking what to do? We aren’t rich and don’t have a car or other means of transport. The treatment costs a lot of money. A rented vehicle was hired to take me to government hospital early morning. Here I am in the last two day and already feeling better. These drips help a lot and provide energy. (IDI, RDW*, *119)*

Furthermore, constitutional symptoms such as weakness, loss of appetite and anemia were often associated with PS among RDW.

### Barriers to Care Seeking for PS

#### Division of labor within the household

Division of labor within the household was identified as a major barrier to care seeking by RDW and their family members. After delivery, majority of new mothers were required to resume household duties and childcare responsibilities, which limited their ability to seek care for themselves.

> *There is no one to work for us. Who will take care of the household chore and look after and tend to children? (IDI, RDW, 275)*.

Generally, women were expected to resume their duties within a few days to a few weeks after delivery. In households with younger female members, the mothers perceived that they would be able to take a break from their duties and seek care. These women also emphasized that no other family members would help with household duties and childcare responsibilities.

#### Decision making at the household level

Through IDIs, RDW highlighted the need for obtaining permission from the primary decision maker prior to seeking care. The senior male of the household was identified as the primary decision maker. The women were not allowed to make the decision to seek care on their own. A family member of RDW shared her insight into the role played by the primary decision maker.

> *Whosoever is elder in the household; father or mother in-law would say that she recently delivered, and she is ill and needs to seek care. If I am elder in the household then I would say this. (IDI, female family member, 127)*.

Additionally, in rural areas, parents-in-law must assess the RDW health situation before allowing her to seek care. In some households, the RDW were able to discuss their health concerns with the primary health decision maker directly or through another family member, who relayed the message to the decision maker. According to RDW, discussing health issues with parents-in-law is not deemed appropriate.

> *I will inform my husband regarding her illness after all she is my daughter. Then we will ask my daughters sister in-law to tell her brother that our daughters sick and needs care. (IDI, female family member, 123)*.

The women also found their husbands unhelpful and unwilling to buy medications and assist them in care seeking. According to family members of RDW, some households prefer the RDW’s family to act as decision makers to care seeking. Majority of the participants expressed that a RDW is taken to hospital only once she is critically ill.

#### Support from family members

Overall, RDW expressed feeling a lack of support from family members when wishing to tend to their healthcare needs and seeking care. The women emphasized feeling depressed and stressed due to the indifference expressed by their husbands and parents-in-law. One RDW described her husband’s reaction to her illness.

> *My husband didn’t ask from me about the illness nor did my in-laws. Rather my husband said, “go die as if I care”. (IDI, RDW, 418)*.

Often, this maltreatment contributes to mothers losing hope of accessing care and getting better. However, many mothers did express the need to improve their health for the sake of their children.

#### Access to transportation

Another major barrier to prompt care seeking was transportation. Often RDW were unable to access care due to a lack of transportation and the vast distant to the health facility. Additionally, the absence of an escort (preferably male) to accompany women to the healthcare providers was a barrier. Participants believed poverty and limited resources were an important factor in preventing them from seeking care. In many situations, TBAs were identified by RDW as being essential in arranging transport to health facilities. TBAs were very resourceful since they were directly in contact with drivers, physicians and other healthcare providers.

#### Limited financial resources

Another critical barrier to care seeking is the lack of financial resources. In Pakistan, public health facilities provide medical care at no cost; however, the patient must pay for the prescription drugs used for treatment. All participants perceived that unemployment and poverty limited the RDW’s ability to seek care from a qualified healthcare provider. They emphasized that families from higher socioeconomic statuses could enjoy the luxury of care seeking and treatment. The RDW with PS interviewed at health facilities also mentioned that lack of funds and transportation are common barriers to accessing care. A RDW with PS shared her perceived barriers to care seeking.

> *These poor people don’t have the money the money which proves a hurdle and they don’t get car for transportation this all add to the worries. (IDI, RDW, 128)*.

All the TBAs mentioned lending money to families to enable them to access tertiary care hospitals. Due to their close links within the community, TBAs were also able to help families access transportation. Often, they have working contracts with healthcare providers and taxi drivers.

#### Alternative sources of care

Alternative sources of care, especially spiritual healers, are a common practice in rural Pakistan. According to the family members of RDW, spiritual healers are very respected by the community. They are an affordable option for financially constrained families. Unfortunately, spiritual healers are known to instill fear and intimidate RDW and their family members. When advised, many RDW mentioned stopping medications previously prescribed by trained healthcare practitioners.

> *I demanded treatment many times however the healer said that stop taking tablets or I will break your neck, so I stopped out of fear. And my mother also stopped my in-laws from taking me to doctors that the healer has said no’ (IDI, RDW, 256)*.

Some RDW and their family members also mentioned seeking care from spiritual healers for fever and anemia. Family members only sought out care from trained healthcare providers once the situation of the RDW had dramatically deteriorated due to fever or anemia.

## Discussion

Maternal health and wellbeing in the postpartum period is directly proportional to the mother’s knowledge of the disease, care seeking behaviours, access to transportation, socioeconomic status and cultural beliefs [8, 14]. Research has shown that behaviour change communication strategies improve care seeking behaviours and access to obstetric care services within households and communities [15, 16]. In our study, the overall care seeking behaviours among RDW for PS depended upon numerous factors such as knowledge of disease, division of labor within the household, the primary decision maker, family support, access to transportation, lack of financial resources and the prominence of spiritual healers in the community. Moreover, as anticipated, the barriers of the Three Delays Model *(socioeconomic and cultural characteristics, accessibility of health facilities* and *quality of care)* were commonly present within the community.

In Matiari, most of the women did not have any knowledge of PS; however, they were able to identify symptoms associated with PS. They were also unaware of the impact of PS on maternal mortality and morbidity. Research suggests that promoting maternal health education, especially the underlying biological cause of PS and related symptoms can help persuade mothers to access appropriate and timely care [5].

In line with previous research, our study Identified that the utilization of healthcare services was not influenced by the women’s perception of illness [17–19]. Although, the severity of the symptoms did influence the RDW’s decision to seek care and patterns of care seeking behaviours. In urban Pakistan, a lack of understanding among women of the symptoms associated with postpartum illnesses was common [5]. Often, these women associated the symptoms of postpartum illnesses with general weakness. A similar lack of understanding related to PS symptoms and incorrect association of symptoms with general weakness was observed among the RDW in rural Pakistan. The RDW also did not consider fever as a dangerous symptom to prompt care seeking.

In Matiari, the status of women plays a major role in their ability to access healthcare. According to the RDW, the primary decision maker, usually an elder male with the highest status within the household would be responsible for deciding whether the women could seek care. This delay in decision making further increases the time in which the women access care as well as their risk of morbidity and mortality [8]. In neighboring countries, the reliance of women on male family members or husbands for decision making pertaining to seeking care is also quite common [20]. During the decision making process, research suggests that other family members do not have the ability to influence or alter the decision.

Accessibility to transportation is a major barrier in lower Income settings, such as rural Pakistan. For women to access public transport, they require a male escort. In the absence of a male escort, women are unable to seek care at health facilities. Similarly, in urban Pakistan, women residing in patrilocal family structures are subjected to male dominance and require a male escort and a face-veil (purdah) when leaving the house [5].

## Strengths & Limitations

The strength of our study is the use of qualitative research methods to identify the barriers to seeking care for PS among RDW. Furthermore, the inclusion of RDW, family members of RDW and healthcare providers’ perspectives strengthens the study. A limitation for our study was the small number of participants interviewed due to logistical constraints. Additionally, due to the study mainly interviewing participants from a rural setting, the findings may not be generalizable at a national level.

## Challenges to data collection

It was difficult to locate women diagnosed with PS in the community and health facilities. During the study recruitment period, only six women with PS were found with four of them consenting to be Interviewed. One reason behind low recruitment may be the dates of the in-depth interviews coinciding with the Holy month of Ramadan. Additionally, the inability of women to seek care due to socioeconomic and cultural barriers may have impacted participant recruitment. Lastly, a low prevalence of PS in the community could have made locating participants challenging.

## Conclusion

The study results have outlined several barriers to care seeking for PS among RDW in rural Pakistan. To improve the patterns of care seeking behaviours for PS, community-based interventions are needed to improve maternal knowledge of PS, address gender inequality, implement an affordable community transport service and enhance TBA’s knowledge and skills to manage PS. The findings of this study will provide sufficient evidence to develop maternal health policies and programs to prevent and manage PS in resource poor settings.

## Data Availability

The data of the study is available upon request keeping in view institutional and ethical polices of the Aga Khan university by emailing to corresponding author.

## Authors’ contributions

SA, UK, AT, SK, FR, FT & FM contributed to manuscript write up and critical revisions. ZAB, LAB AL, provided critical technical inputs. SBS reviewed final manuscript. All authors read and approved the final manuscript.

## Acknowledgements

The authors gracefully acknowledge the contribution of all field team members particularly Mr Syed Shujaat Hussain Zaidi & Dr Sheraz Memon for their hard work and support. We would also like to thank all study participants

## Data Sharing

The data of the study is available upon request keeping in view Institutional and ethical polices of the Aga Khan university by emailing to corresponding author.

## References

1. Madhudas C, Khurshid F, Sirichand P. Maternal Morbidity and Mortality Associated with Puerperal Sepsis. JLUMHS. 2011 Dec; 10(3).

2. Dillen JV, Zwart J, Schutte J, Roosmalen JV. Maternal sepsis: epidemiology, etiology and outcome. Curr Opin Infect Dis. 2010; 23:249–254.

3. Hogan MC, Foreman KJ, Naghavi M, Ahn SY, Wang M, Makela SM et al., Maternal mortality for 181 countries, 1980-2008: a systematic analysis of progress towards Millennium Development Goal 5. Lancet. 2010 Apr; 375 (9726): 1609–23.

4. World Health Organization.The global burden of disease: 2004 update [Internet]. Geneva: World Health Organization; 2008 [cited 2016 May 05]. Available from: http://www.who.int/healthinfo/global_burden_disease/GBD_report_2004update_full.pdf?ua=1

5. Fikree FF, An T, Durocher JM, Rahbar MH. Health service utilization for perceived postpartum morbidity among poor women living in Karachi. Social Sciences and Medicine. 2004 Aug; 59(4): 681–694.

6. National Institute of Population Studies (NIPS), ICF International. Pakistan Demographic and Health Survey 2012-13. Islamabad, Pakistan, and Calverton, Maryland, USA: NIPS and ICF International; 2013 [cited 2016 May 05]. Available from: http://www.nips.org.pk/abstract_files/PDHS%20Final%20Report%20as%20of%20Jan%2022-2014.pdf

7. Moran AC, Winch PJ, Sultana N, Kalim N, Afzal KM, Koblinsky M et al. Patterns of maternal care seeking behaviours in rural Bangladesh. Trop Med Int Health. 2007 Jul; 12(7): 823–32.

8. Thaddeus S, Maine D. Too far to walk: Maternal Mortality in Context. Social Sciences and Medicine. 1994 Apr; 38(8):1091–110.

9. Maternity Worldwide. The Three Delays Model and our Integrated Approach [Internet]. Brighton: Maternity Worldwide; 2015 [cited 2016 May 03]. Available from: http://www.maternityworldwide.org/what-we-do/three-delavs-model/

10. Calvello EJ, Skog AP, Tenner AG, Wallis LA. Applying the lessons of maternal mortality reduction to global emergency health. Bull World Health Organ. 2015 Jun; 93 (6).

11. Papali A, McCurdy MT, Calvello EJ. A “three delays” model for severe sepsis in resource-limited countries. Journal of Critical Care. 2015; 30: 861.e9–861.e14.

12. National Institute of Population Research and Training (NIPORT), Mitra and Associates, ORC Macro. Bangladesh Demographic and Health Survey 1999-2000. Dhaka, Bangladesh and Calverton, MD, USA: NIPORT, Mitra and Associates and ORC Macro; 2001 [cited 2016 May 03]. Available from: https://dhsprogram.com/pubs/pdf/FR119/FR119.pdf

13. Stewart MK, Stanton CK and Ahmed O. Demographic and Health Surveys Comparative Studies: Maternal Health Care. Calverton, MD, USA: Macro International Inc; 1997 Sept [cited 2016 May 03]. Available from: http://dhsprogram.com/pubs/pdf/CS25/CS25.pdf

14. McCarthy J, Maine D. A framework for analyzing the determinants of maternal mortality. Stud Fam Plann. 1992 Feb; 23 (1): 23–33.

15. Koblinsky, M. Improving obstetrical and neonatal management: Lessons from Guatemala. MotherCare Matters. 1996 Aug; 5(4): 1–3.

16. Koblinsky M, Favin M, Kureshy N, Elder L. Behavioral dimensions of maternal health and survival. A consultative forum. MotherCare Matters. 2000 Sep; 9(3): 1–19.

17. Bhatia JC, Cleland J. Obstetric morbidity in South India: Results from a community survey. Soc Sci Med. 1996 Nov; 43 (10): 1507–1516.

18. Stewart MK, Stanton CK, Festin M, Jacobson N. Issues in measuring maternal morbidity: Lessons from the Philippines Safe Motherhood Project. Stud Fam Plann. 1996 Feb; 27(1): 29–35.

19. Kureshy N. Safe Motherhood Project (Korangi 8, Karachi): Formative Research Report. Karachi, Pakistan: The Aga Khan University and MotherCare/John Snow Inc; 1998. Pakistan.

20. Gupta MD. Selective Discrimination Against Female Children in Rural Punjab, India. Population and Development Review. 1987 Mar; 13(1): 77–100.

